# Modelling the impact of contact tracing of symptomatic individuals on the COVID-19 epidemic

**DOI:** 10.1101/2021.02.05.21251222

**Authors:** Marcos Amaku, Dimas Tadeu Covas, Francisco Antonio Bezerra Coutinho, Raymundo Soares Azevedo, Eduardo Massad

## Abstract

**OBJECTIVES:** With declining numbers of COVID-19 cases in the State of São Paulo, Brazil, social distancing measures were gradually being lifted. The risk of an increase in the number of cases, however, cannot be overlooked. Even with the adoption of non-pharmaceutical interventions, such as restrictions on mass gatherings, wearing masks, and complete or partial closure of schools, other public health measures may help to control the epidemic. We aimed to evaluate the impact of the contact tracing of symptomatic individuals on the COVID-19 epidemic regardless of the use of diagnostic testing.

**METHODS:** We developed a mathematical model that includes isolation of symptomatic individuals and tracing of contacts to assess the effects of the contact tracing of symptomatic individuals on the COVID-19 epidemic in the State of São Paulo.

**RESULTS:** For a selection efficacy (proportion of isolated contacts who are infected) of 80%, cases and deaths may be reduced by 80% after 60 days when 5000 symptomatic individuals are isolated per day, each of them together with 10 contacts. On the other hand, for a selection efficacy of 20%, the number of cases and deaths may be reduced by approximately 40% and 50%, respectively, compared with the scenario in which no contact tracing strategy is performed.

**CONCLUSION:** Contact tracing of symptomatic individuals may be a potential alternative strategy when the number of diagnostic tests available is not sufficient for a massive testing strategy.

## Introduction

The first case of COVID-19 in Brazil was reported on 26 February 2020 in the State of São Paulo, the most populous Brazilian state with 44,639,899 inhabitants (1). Genome sequencing coupled with phylogenetic analyses corroborate multiple importations of the virus from Italy followed by local spread (2). As of 30 October 2020, 1,113,788 cases and 39,255 deaths were reported in Brazil (3), the largest numbers in Latin America (4).

Isolation, quarantine, social distancing and community containment are important non-pharmaceutical public health interventions to control the explosive growth of COVID-19 (5). Liberal testing, followed by contact tracing and isolation of all test positive persons have direct and clear benefits (6).

The World Health Organization recommends a combination of rapid diagnosis, immediate isolation of cases, rigorous tracking and precautionary self-isolation of close contacts (6). In a previous paper, we analyzed the impact and costs of test-trace-quarantine strategies (7). Here we set out to model the effects of a contact tracing strategy of symptomatic individuals to control the COVID-19 spread regardless of the use of diagnostic testing. This may be an alternative strategy for places with limited availability of diagnostic tests.

## METHODS

### The model

The model is based on a modified version of the SEIR model (7,8) and considers that the population at time *t* is divided into several compartments, namely: susceptible individuals, *S*(*t*); isolated susceptible individuals, *Q*_*S*_(*t*); susceptible individuals previously isolated,; exposed individuals, *E*(*t*); asymptomatic/oligosymptomatic individuals, *A*(*t*); symptomatic individuals, *I*(*t*); isolated infected individuals, *Q*(*t*); hospitalized individuals, *H*(*t*); individuals with severe disease hospitalized in intensive care units (ICU), *G*(*t*); and recovered individuals, *R*(*t*).

A schematic representation of the model is shown in Figure 1.

**Figure 1.**
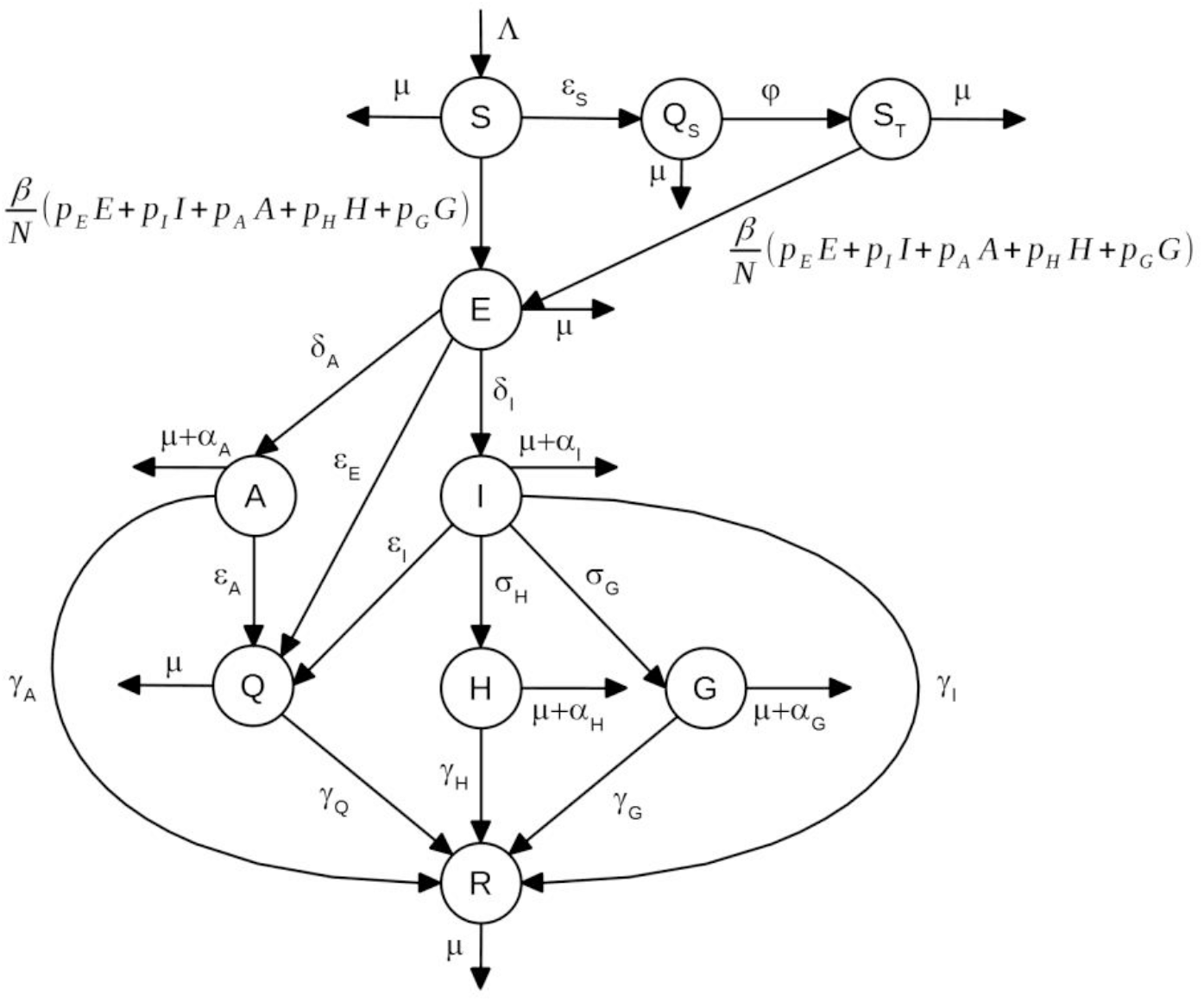
Schematic representation of the model compartments.

The dynamics of individuals between compartments may be described as follows:

i. Susceptible individuals, *S*(*t*), grow with a birth rate Λ(*t*), either may acquire the infection with contact rate *β*, or be isolated with constant rate *ε*_*S*_ (i.e., *ε*_*S*_ individuals isolated per unit time).
ii. Isolated susceptible individuals,. *Q*_*S*_ (*t*), after a period of 1/ *φ*, are moved to the compartment *S*_*T*_(*t*).
iii. Once infected, the susceptible *S*(*t*) and *S*_*T*_(*t*) individuals move to the state of exposed individuals, denoted *E*(*t*).
iv. Exposed individuals may evolve to symptomatic individuals, *I*(*t*), with rate *δ* _*I*_, or evolve to asymptomatic/oligosymptomatic individuals, denoted and may be isolated with constant rate *ε*_*E*_. *A*(*t*), with rate *δ*_*A*_, and may be isolated with constant rate *ε*_*E*_.
v. Infectious individuals, *I*(*t*), may either evolve to one of two states, hospitalized individuals denoted *H*(*t*), with rate *σ*_*H*_, or to a state in which individuals develop severe disease and are admitted to intensive care units, denoted *G*(*t*), with rate *ε*_*G*_. Infectious individuals, *I*(*t*), may be isolated with constant rate *ε*_*I*_ and may also die by the disease, with rate *α*_*I*_
vi. Asymptomatic individuals *A*(*t*) may be isolated with constant rate *ε*_*A*_.
vii. Individuals in the states *A*(*t*), *H*(*t*), and *G*(*t*) may die by the disease, with rates *α*_*A*_, *α*_*H*_, and *α*_*G*_, respectively.
viii. All individuals who acquired the infection and who did not die by the disease, recover to a new state, denoted. *R*(*t*), with rates *γ*_*I*_, *γ*_*A*_, *γ*_*H*_ and *γ*_*G*_, as depicted in Figure 1
ix. Isolated infected individuals are moved to a state denoted *Q*(*t*). Since these individuals are isolated from the rest of the population, they do not transmit the virus and will eventually recover from the infection, with rate *γ*_*Q*_.
x. All individuals may die by natural causes with rate *μ*.
xi. We assumed that the population birth rate ^(*t*) is equal to the natural mortality of the population, disregarding the disease-induced mortality.
xii. The fractions *p*_*E*_, *p*_*I*_, *p*_*A*_, *p*_*H*_, and *p*_*G*_ of exposed, symptomatic, asymptomatic, hospitalized and severe (ICU patients) individuals can transmit the infection.

The following set of differential equations describes the model dynamics.

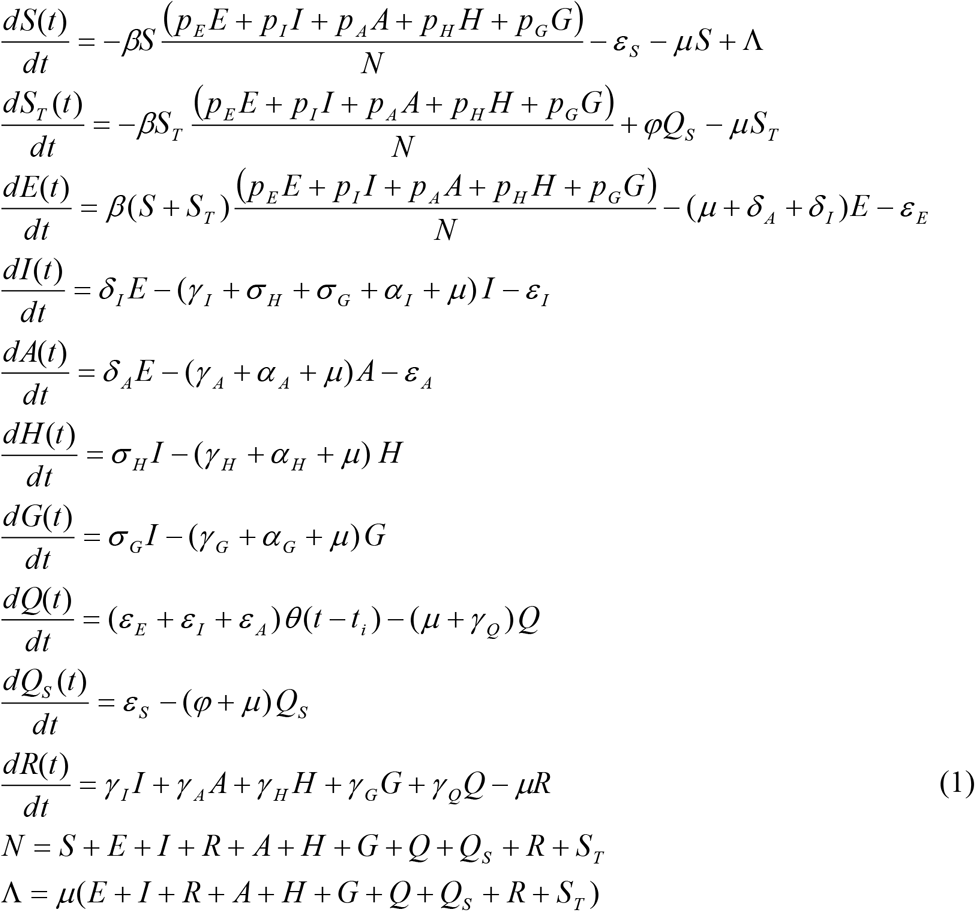

The basic reproduction number of system (1) is given by

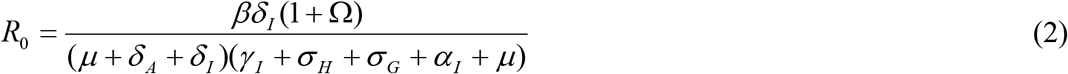

where

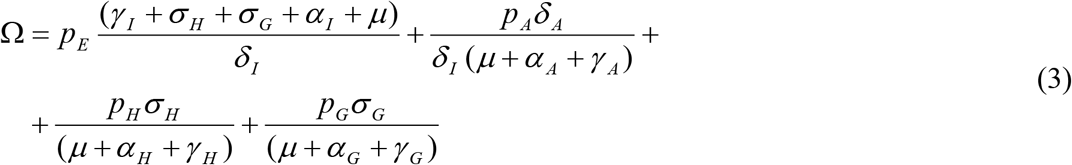

The incidence of infection is given by:

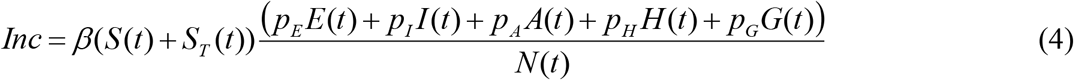

The total number of reported cases is obtained by multiplying the number of infected individuals by a notification ratio *K*(*t*):

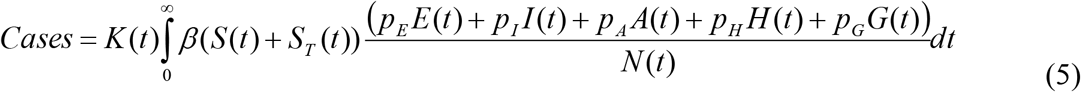

The total number of COVID-19-related deaths is given by:

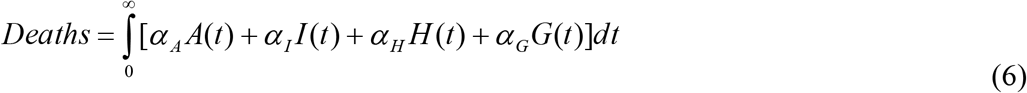

Finally, the total number of isolated individuals is given by:

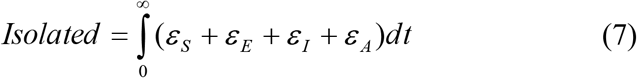

If the number of symptomatic individuals in a certain time interval Δ*t* is less Than *ε*_*I*_ Δ*t*, only the available symptomatic individuals are isolated together with their contacts. A similar procedure is adopted for the number of susceptible and asymptomatic individuals in the compartments *S, E* or *I* when they are below *ε*_*S*_ Δ*t, ε*_*E*_ Δ*t* or *ε*_*A*_Δ*t*, respectively.

### Fitting procedure

We used the fitting procedure proposed by Amaku et al. (7) and described as follows.

Data on the cumulative number of reported cases and deaths were obtained from Seade (Fundação Sistema Estadual de Análise de Dados do Estado de São Paulo). Data on the number of ICU patients were obtained from SIMI (Sistema de Monitoramento Inteligente do Estado de São Paulo). A fitting procedure based on the Levenberg-Marquardt non-linear least-squares algorithm was used to fit the model’s parameters simultaneously to the data on cases, deaths and ICU patients. We used the R package minpack.lm (Elzhov et al., 2016).

We assumed that the potentially infective contact rate, the notification ratio, and the ICU admission rate change every 10 days.

The parameter values used are shown in Table 1.

**Table 1.**
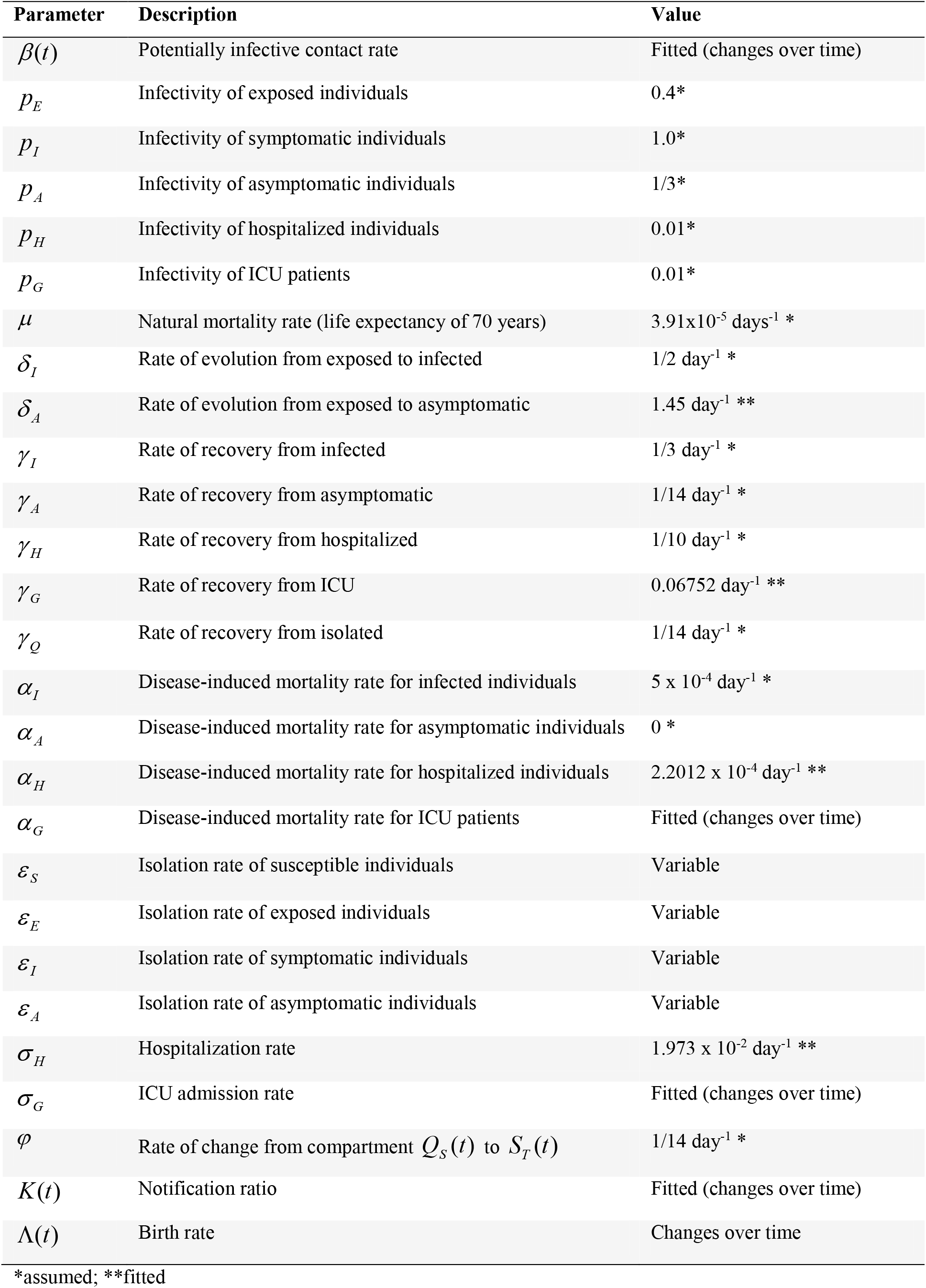
Parameters used in the model.

| Parameter | Description | Value |
| --- | --- | --- |
| $\beta(t)$ | Potentially infective contact rate | Fitted (changes over time) |
| $p_E$ | Infectivity of exposed individuals | 0.4* |
| $p_I$ | Infectivity of symptomatic individuals | 1.0* |
| $p_A$ | Infectivity of asymptomatic individuals | 1/3* |
| $p_H$ | Infectivity of hospitalized individuals | 0.01* |
| $p_G$ | Infectivity of ICU patients | 0.01* |
| $\mu$ | Natural mortality rate (life expectancy of 70 years) | $3.91 \times 10^{-5} \text{ days}^{-1}$ * |
| $\delta_I$ | Rate of evolution from exposed to infected | $1/2 \text{ day}^{-1}$ * |
| $\delta_A$ | Rate of evolution from exposed to asymptomatic | $1.45 \text{ day}^{-1}$ ** |
| $\gamma_I$ | Rate of recovery from infected | $1/3 \text{ day}^{-1}$ * |
| $\gamma_A$ | Rate of recovery from asymptomatic | $1/14 \text{ day}^{-1}$ * |
| $\gamma_H$ | Rate of recovery from hospitalized | $1/10 \text{ day}^{-1}$ * |
| $\gamma_G$ | Rate of recovery from ICU | $0.06752 \text{ day}^{-1}$ ** |
| $\gamma_Q$ | Rate of recovery from isolated | $1/14 \text{ day}^{-1}$ * |
| $\alpha_I$ | Disease-induced mortality rate for infected individuals | $5 \times 10^{-4} \text{ day}^{-1}$ * |
| $\alpha_A$ | Disease-induced mortality rate for asymptomatic individuals | 0 * |
| $\alpha_H$ | Disease-induced mortality rate for hospitalized individuals | $2.2012 \times 10^{-4} \text{ day}^{-1}$ ** |
| $\alpha_G$ | Disease-induced mortality rate for ICU patients | Fitted (changes over time) |
| $\varepsilon_S$ | Isolation rate of susceptible individuals | Variable |
| $\varepsilon_E$ | Isolation rate of exposed individuals | Variable |
| $\varepsilon_I$ | Isolation rate of symptomatic individuals | Variable |
| $\varepsilon_A$ | Isolation rate of asymptomatic individuals | Variable |
| $\sigma_H$ | Hospitalization rate | $1.973 \times 10^{-2} \text{ day}^{-1}$ ** |
| $\sigma_G$ | ICU admission rate | Fitted (changes over time) |
| $\varphi$ | Rate of change from compartment $Q_S(t)$ to $S_T(t)$ | $1/14 \text{ day}^{-1}$ * |
| $K(t)$ | Notification ratio | Fitted (changes over time) |
| $\Lambda(t)$ | Birth rate | Changes over time |

\*assumed; \*\*fitted

Model projections for future dates were obtained by keeping fixed the fitted values of the parameters from the last date observed in the data.

### The contact tracing (CT) strategy

A number of symptomatic individuals are isolated per unit time. *c* contacts of each symptomatic individual are also isolated. We varied both *ε*_*I*_ and *c*. Isolated individuals remain in isolation during 14 days.

Assuming that a fraction of the isolated contacts may be susceptible or recovered, we define a selection efficacy as the proportion of isolated contacts who are infected (asymptomatic or symptomatic individuals).

We calculated the efficacy of the CT strategy subtracting from 1 the result of the division of the cumulative number of cases by the number of cases in the baseline scenario in which CT is not performed.

We assumed an initial condition with 15%, 83% and 2% of recovered, susceptible and infected individuals. These estimates are consistent with the model projections for the beginning of August, 2020 in the State of São Paulo. Among the infected individuals, an asymptomatic-to-symptomatic ratio of 5 and a ratio of asymptomatic-to-exposed (from the compartments A and E) of 9 were assumed.

### Sensitivity analysis

A sensitivity analysis was performed using a Monte Carlo method to sample parameter values and a PRCC (Partial Rank Correlation Coefficient) estimation to measure the strength of association between an input parameter and an output variable after the linear effects on the output variable of the remaining inputs are discounted (9,10). Parameter values were sampled using a Monte Carlo sampling method assuming a uniform distribution for each parameter. The following input parameters were included in the analysis: proportions of susceptible (*f*_*S*_), infected (*f*_*I*_), and recovered (*f*_*R*_) individuals in the initial condition; number of symptomatic individuals isolated (*ε*_*I*_) per unit time; number of contacts (*c*) of each symptomatic individual isolated; the selection efficacy (*eff*); and the asymptomatic-to-symptomatic ratio (*r*_*AS*_). The ranges of parameter values used in the sensitivity analysis are shown in Table 2. The cumulative number of cases after 60 days was used as the output variable.

**Table 2.** Ranges of parameter values used in the sensitivity analysis. The output variable was the cumulative number of cases after 60 days and the input parameters are described in the table. Parameter values were sampled using a Monte Carlo sampling method assuming a uniform distribution.

| Input parameter | Description | Range |
| --- | --- | --- |
| $f_s$ | Proportion of susceptible individuals in the initial condition | Uniform (min=0.5, max=0.9) |
| $f_I$ | Proportion of infected individuals in the initial condition | Uniform (min=0.005, max=0.02) |
| $f_R$ | Proportion of recovered individuals in the initial condition | $1 - f_s - f_I$ |
| $\varepsilon_I$ | Number of symptomatic individuals isolated per day | Uniform (min=500, max=5000) |
| $c$ | Number of contacts | Uniform (min=5, max=10) |
| $eff$ | Selection efficacy | Uniform (min=0.2, max=0.8) |
| $r_{AS}$ | Asymptomatic-to-symptomatic ratio | Uniform (min=1/5, max=5/1) |

## RESULTS

We fitted the model parameters simultaneously to the data of cumulative number of reported cases, deaths and the number of ICU patients (Figure 2) for the state of São Paulo until July 18, 2020. To estimate a 95% probability interval (shaded area in Figure 2), we assumed a normal distribution for the contact rate with a standard deviation of 1.0%.

**Figure 2.**
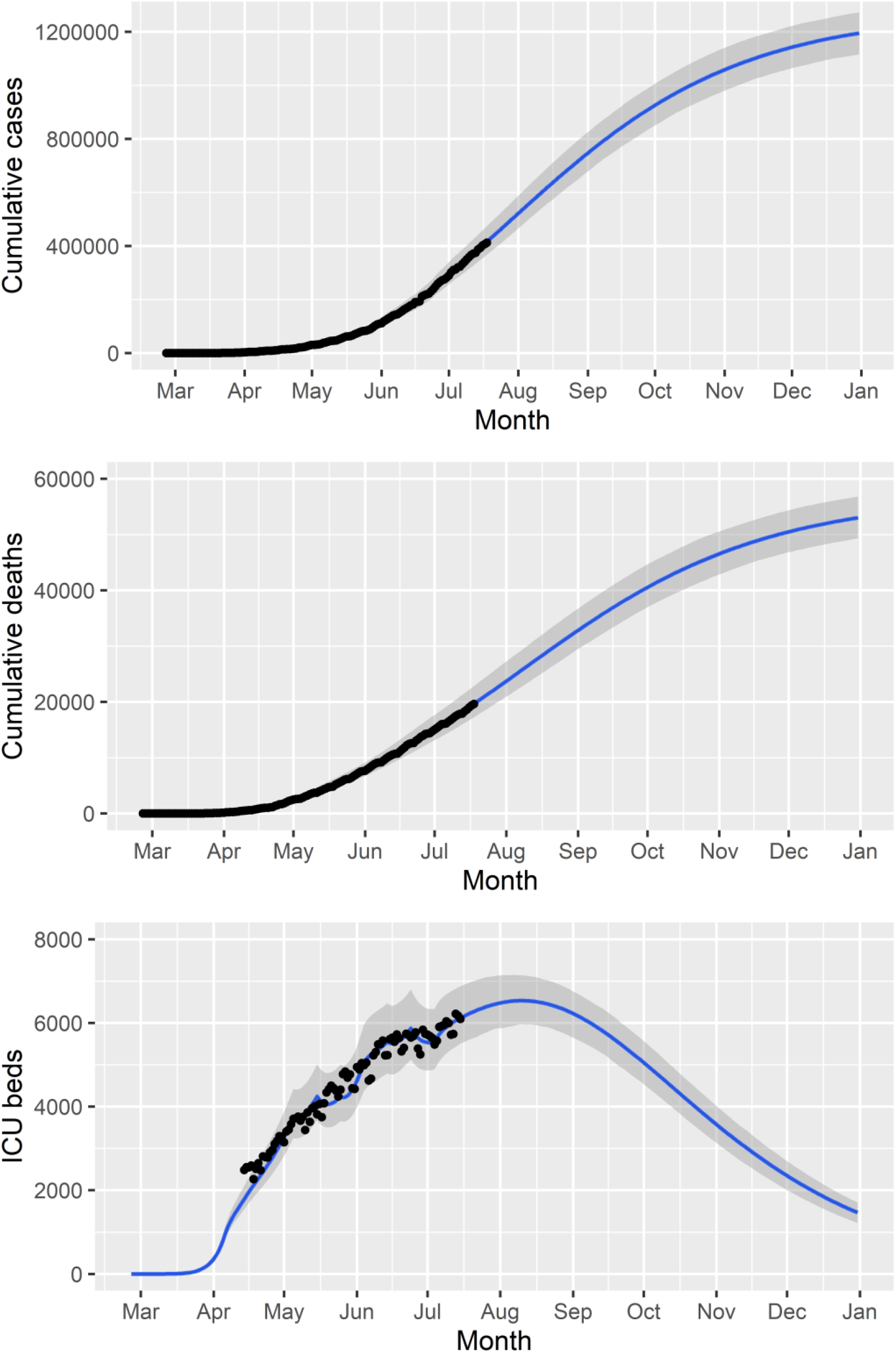
Cumulative number of reported cases and deaths, and number of ICU patients (black dots) and the corresponding fitted model (blue lines). The solid lines and shaded area correspond, respectively, to median values and 95% probability intervals.

The cumulative number of cases and deaths over time for different numbers (1000, 3000 or 5000) of symptomatic individuals isolated per day and their contacts (5 or 10 contacts per symptomatic individuals) are shown in Figures 3 and 4, respectively. Selection efficacies of 20% and 80% were taken into account. The solid line shows the results when no CT strategy was used. The efficacy of the CT strategies and the number of isolated individuals are shown in Figures 5 and 6, respectively.

**Figure 3.**
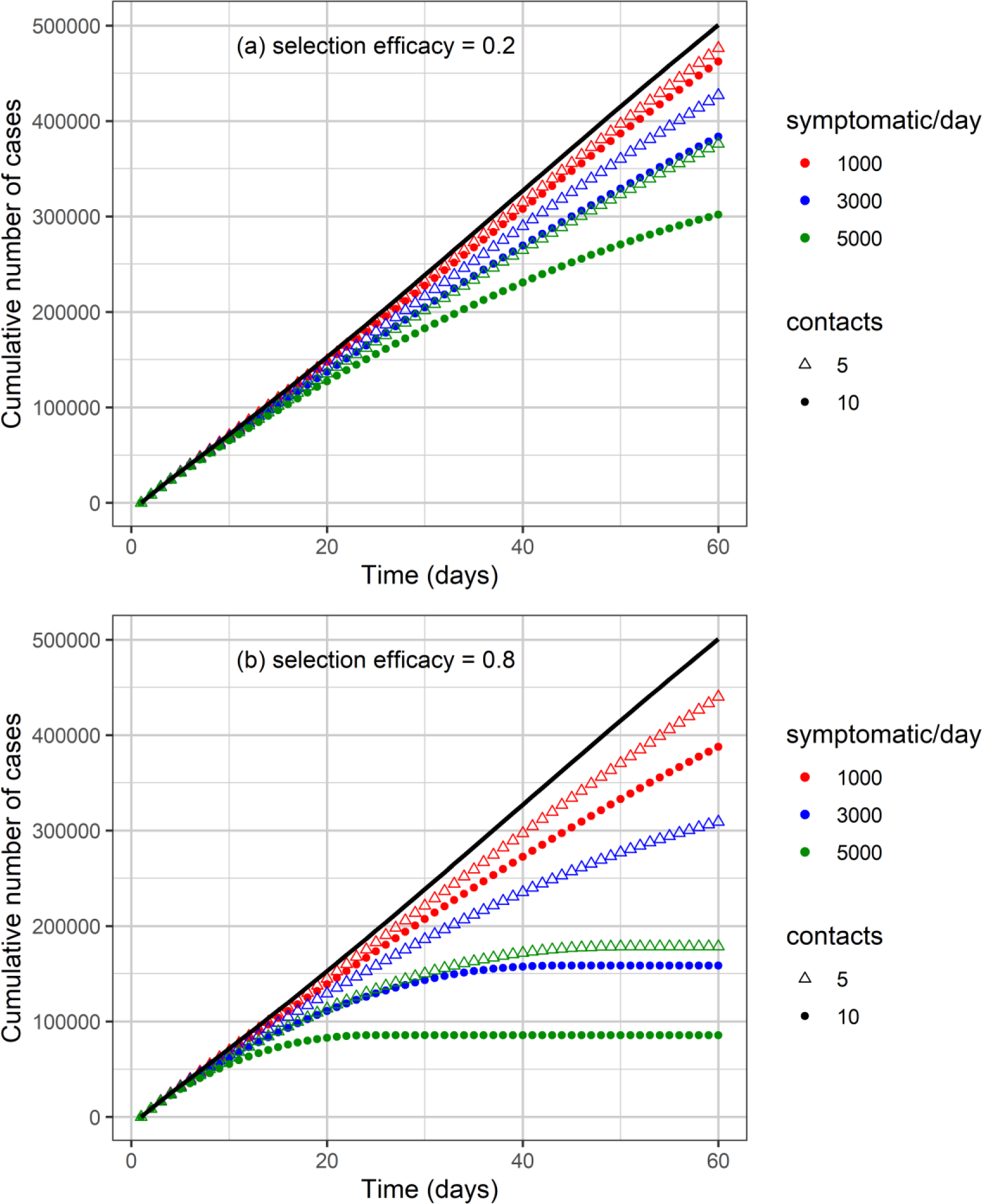
Cumulative number of cases as a function of time for different numbers of isolated symptomatic individuals per day, isolated contacts and selection efficacy of (a) 20% and (b) 80%. The solid black line shows the effect that would be observed if no isolation strategy is used.

**Figure 4.**
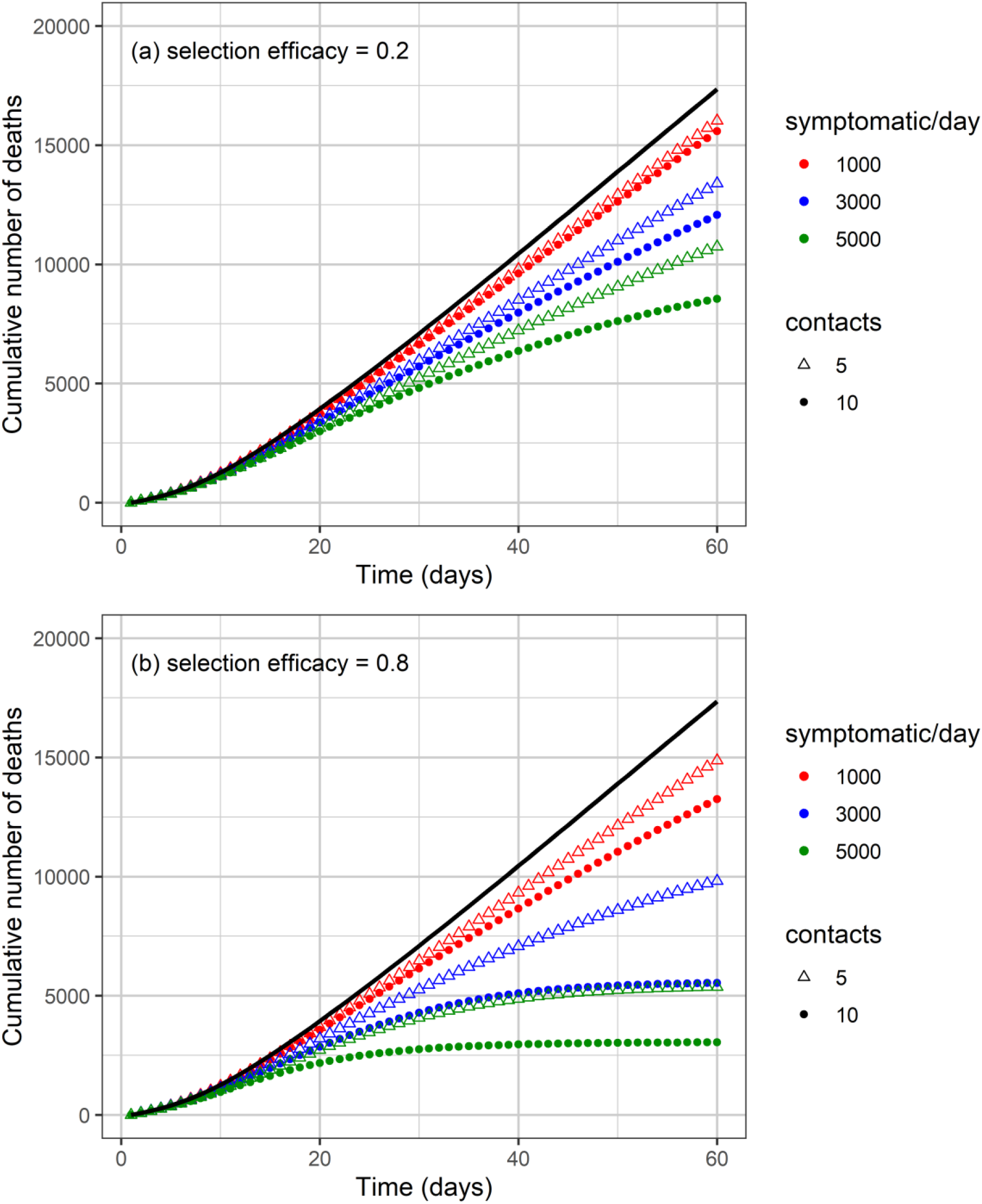
Cumulative number of deaths as a function of time for different numbers of isolated symptomatic individuals per day, isolated contacts and selection efficacy of (a) 20% and (b) 80%. The solid black line shows the effect that would be observed if no isolation strategy is used.

**Figure 5.**
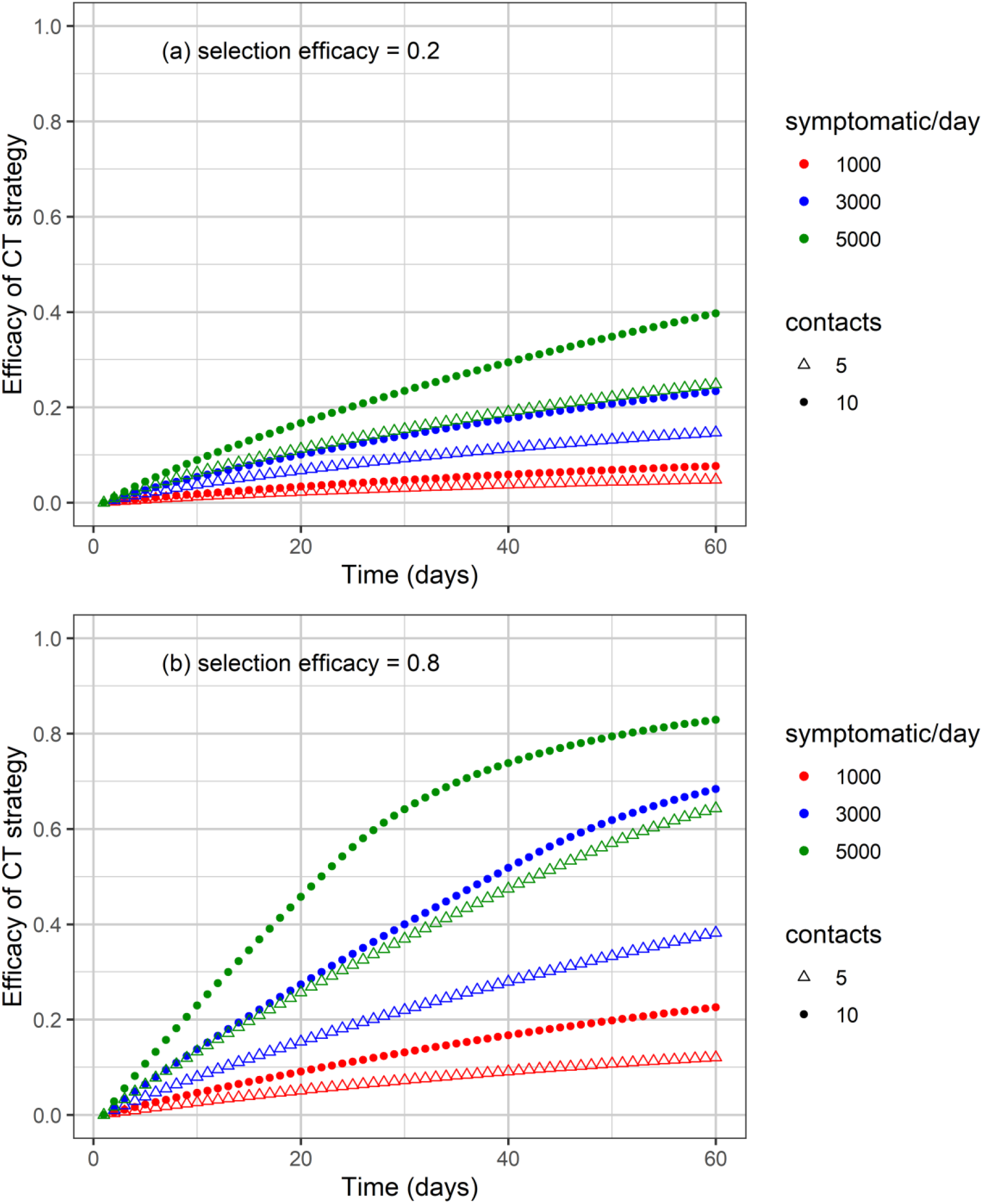
Efficacy of the CT strategy, defined as 1 minus the ratio of the number of cases under a CT strategy divided by the number of cases without CT strategy, as a function of time for different combinations of symptomatic individuals isolated per day, number of isolated contacts and selection efficacy of (a) 20% and (b) 80%.

**Figure 6.**
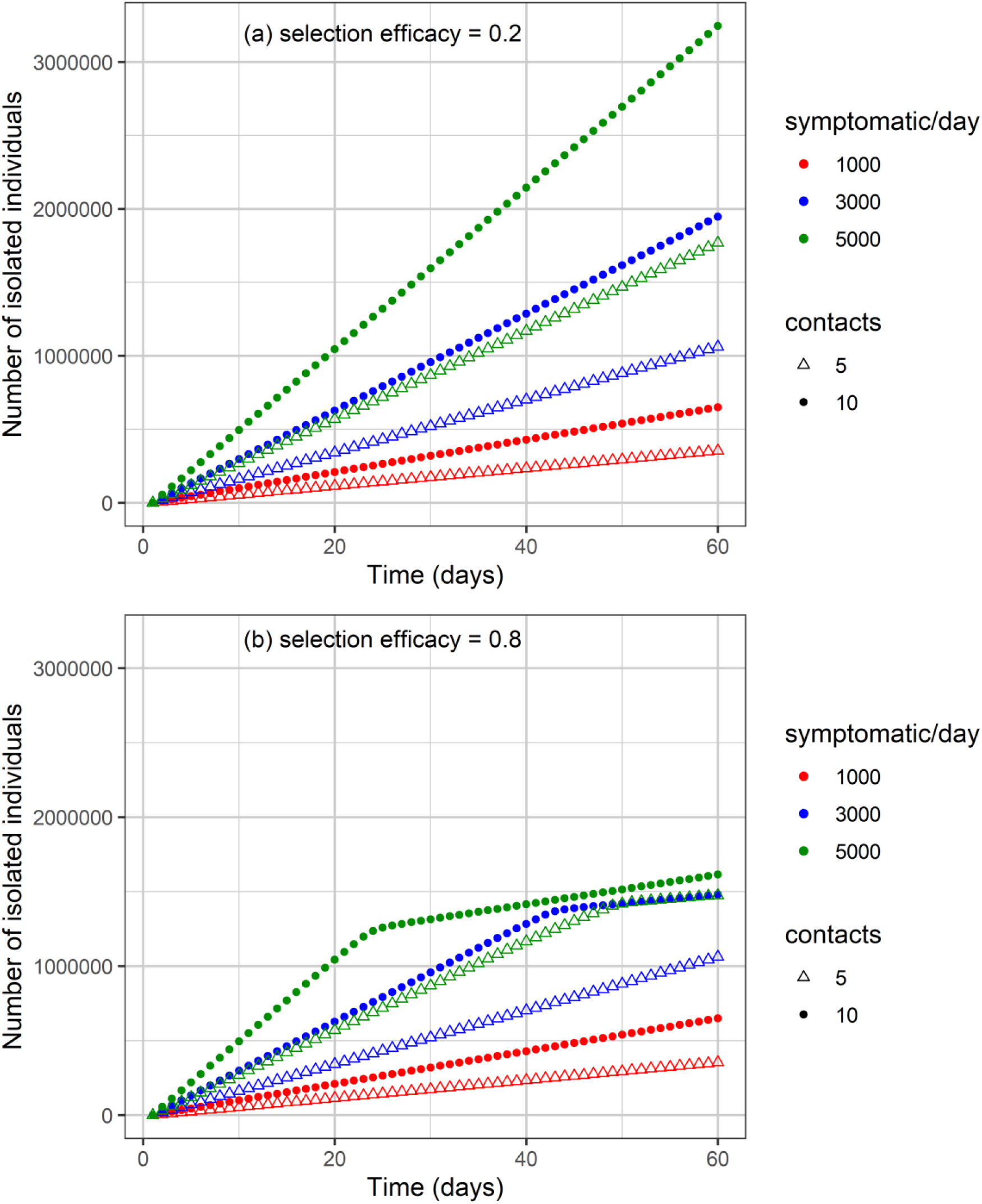
Cumulative number of isolated individuals as a function of time for different combinations of isolated symptomatic individuals per day, isolated contacts and selection efficacy of (a) 20% and (b) 80%.

The higher the number of symptomatic individuals isolated per day, the lower the cumulative number of cases and deaths (Figures 3 and 4). For instance, when 5000 symptomatic individuals are isolated per day, each of them together with 10 contacts, the number of cases and deaths are reduced by approximately 40% and 50%, respectively, compared with the scenario in which the CT strategy is not performed, for a selection efficacy of 20% and 60 days since the beginning of the strategy implementation. For the selection efficacy of 80%, cases and deaths are reduced by 80% approximately.

As the calculation of the efficacy of the CT strategy is based on the reduction in the number of cases, for the scenarios described in the previous example, the efficacy of the CT strategy is 40% for the selection efficacy of 20% (Figure 5). For the selection efficacy of 80%, the efficacy of the CT strategy is 82% approximately.

Regarding the number of isolated individuals, when the selection efficacy is low (20%), the number of isolated individuals may be as high as 3.2 million isolated individuals after 60 days for the strategy with 5000 symptomatic individuals isolated per day together with 10 contacts for each individual (Figure 6). On the other hand, when the selection efficacy is high (80%), about 1.6 million individuals were isolated after 60 days.

PRCC values are shown in Figure 7. The sign of the PRCC is related to the qualitative relationship between the input parameter and the output variable (number of cumulative cases). The number of cumulative cases decreases as the number of symptomatic individuals isolated (*ε*_*I*_), the selection efficacy (*eff*) and the number of contacts (*c*) increase, thus the PRCC values are negative. The positive values of the PRCC for the initial proportion of infected individuals (*f*_*I*_) and the asymptomatic-to-symptomatic ratio (*r*_*AS*_) imply that, when these parameters increase, the number of cumulative cases also increase. The PRCC values for the initial proportion of susceptible (*f*_*S*_) and recovered (*f*_*R*_) individuals are positive but closer to zero (low correlation) when these two initial condition values are in the ranges shown in Table 2.

**Figure 7.**
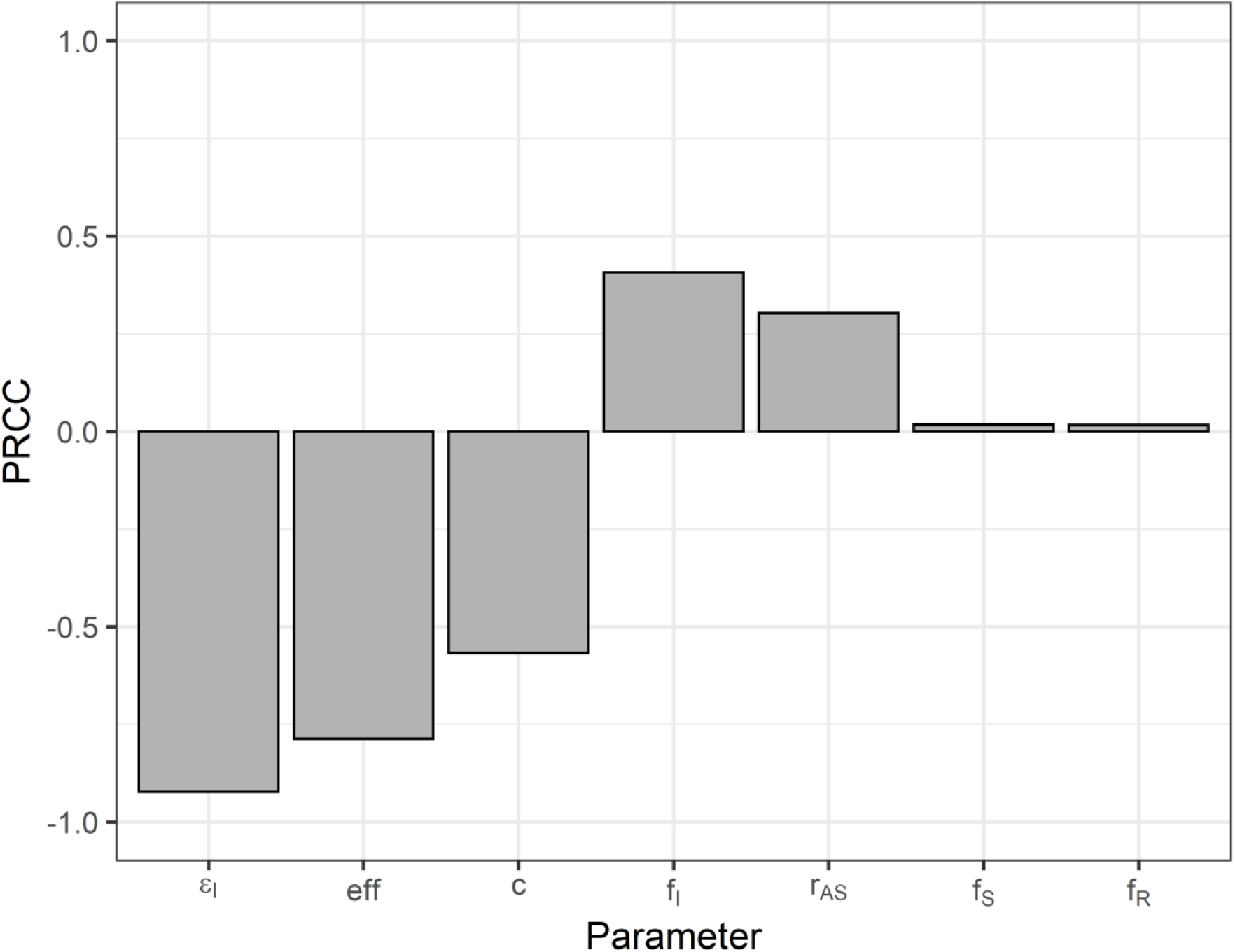
Partial rank correlation coefficients for the number of cumulative cases as output variable and the following input variables: number of symptomatic individuals isolated (*ε*_*I*_), selection efficacy (*eff*), number of contacts (*c*), initial proportion of infected (*f*_*I*_), susceptible (*f*_*S*_) and recovered (*f*_*R*_) individuals, and asymptomatic-to-symptomatic ratio (*r*_*AS*_).

## DISCUSSION

We modelled the impact of a strategy based on contact tracing of symptomatic individuals on the COVID-19 epidemic in the State of São Paulo, Brazil. This strategy has lower costs when compared to a test-trace-and-quarantine strategy (7). It may be a potential alternative strategy when the number of diagnostic tests available is not sufficient for a massive testing strategy.

In the sensitivity analysis, we observed that the reduction in the number of cumulative cases was more sensitive to the number of symptomatic individuals isolated, the selection efficacy, and the number of contacts, in decreasing order of the PRCC. The increase in the number of isolated symptomatic individuals and their contacts poses logistical challenges and associated costs. These costs, however, are likely to be lower than the costs of a test, trace and quarantine strategy (7).

The higher the selection efficacy, the higher the efficacy of the CT strategy (Figure 5). The use of high performance diagnostic tests would likely increase the selection efficacy. However, without the use of diagnostic tests, one could think that tracing of close contacts of symptomatic individuals, such as household members or coworkers, would probably increase the selection efficacy, thus increasing the overall efficacy of the CT strategy.

Optimising tracing coverage and minimizing tracing delays, for instance with app-based technology, further enhance contact tracing effectiveness, as pointed out by Kretzschmar et al. (11). As discussed by Bilinski et al. (12), the benefits of contact tracing depend on adherence to isolation and quarantine among individuals who are traced. The adherence may be enhanced by measures such as out-of-home accommodations, income replacement, and social supports (12).

A limitation of this analysis is that we are assuming that the isolated symptomatic individuals are infected by SARS-CoV-2 and not by other virus that could cause similar symptoms. This limitation, however, would be less important in a scenario in which a substantial part of the respiratory infections are caused by SARS-CoV-2. Nevertheless, one could interpret the number of isolated symptomatic individuals as an effective number of individuals infected by SARS-CoV-2 who should be isolated in order to observe the outcomes of the model.

## CONCLUSION

We evaluated the impact of contact tracing of symptomatic individuals and their contacts on the number of cases and deaths related to COVID-19. Depending on the number of symptomatic individuals isolated per day and also on the efficacy of selecting infected (asymptomatic) contacts to be isolated, the overall efficacy of the contact tracing strategy can be high. For instance, for a selection efficacy of 80%, cases and deaths may be reduced by 80% after 60 days when 5000 symptomatic individuals are isolated per day, each of them together with 10 contacts. On the other hand, for a selection efficacy of 20%, the number of cases and deaths may be reduced by approximately 40% and 50%, respectively, compared with the scenario in which no contact tracing strategy is performed. Thus, contact tracing of symptomatic individuals may be a potential alternative strategy when the number of diagnostic tests available is not sufficient for a massive testing strategy.

## Data Availability

The data on COVID-19 cases and deaths are available from https://www.seade.gov.br/

## ACKNOWLEDGMENTS

This work was partially supported by LIM01-HFMUSP, CNPq, FAPESP and Fundação Butantan.

## AUTHOR CONTRIBUTIONS

Amaku M, Covas DT, Coutinho FAB, Azevedo RS and Massad E participated in the modelling design, discussion of the subject, writing and revision of the manuscript.

## Notes

### Competing Interest Statement

The authors have declared no competing interest.

### Funding Statement

This work was partially supported by LIM01-HFMUSP, CNPq, FAPESP and Fundacao Butantan.

### Author Declarations

Ethical approval. Not applicable.

